# Beyond AI Psychosis and Sycophancy: Structural Drift as a System-Level Safety Failure

**DOI:** 10.64898/2026.03.19.26346371

**Authors:** Jasmine E. Kim, Erik Holbrook, Jonathan D. Hron, Chase R. Parsons

## Abstract

**Background:** Conversational AI safety systems are primarily evaluated using message-level content monitoring, which assesses inputs and outputs in isolation. This message-by-message approach can miss interaction-level risks that emerge over extended conversations, including patterns discussed in reports of “AI psychosis.” Critically, by the time users express overt psychosis-spectrum content, opportunities for intervention may be limited.

**Objective:** We investigated whether LLM responses gradually expand and connect interpretations beyond the user’s original concerns, a process we term *structural drift*. We also tested whether this drift can be detected early and automatically.

**Methods:** We developed an automated, LLM-adapted rubric-based prompt for seven domains of anomalous (psychosis-spectrum) experience, derived from phenomenological psychiatry to capture subtle shifts in subjective interpretation. In Part 1, we evaluated the rubric using gold-standard text excerpts (N = 484) adapted from clinically validated qualitative instruments. In Part 2, we analyzed 1,290 user-LLM response exchanges from 7 dialogues, using 3 different LLMs (5 repeats each), to measure (i) domain amplification (increasing score within a domain) and (ii) domain expansion (new domains appearing over time).

**Results:** Automated scoring showed strong agreement with gold-standard excerpts (domain accuracy 82.7-98.9%; exact 0-3 agreement 63.6-82.7%). Across dialogues, we observed significant amplification in four domains (*p* < .05; *d* = 0.14-0.46) and domain expansion in 83.8% of dialogues (88/105; *p* < .001).

**Conclusions:** AI responses can systematically expand and intensify users’ descriptions beyond their initial input. Taken together with the predictive-processing accounts of psychosis, the exposure itself may reinforce maladaptive inferences. Because drift is detectable from ordinary dialogue without clinical-style probing, this structural drift detection may support scalable, real-time monitoring for emerging risks before overt escalation.

## INTRODUCTION

There are increasing accounts of AI users experiencing psychological harms from prolonged use, with firsthand accounts and case studies documenting these problems.^1–4^ Yet the mechanism remains unclear,^5–7^ leaving safety systems unable to detect these risks. This raises a foundational challenge: how can AI systems recognize when continued interaction is doing more harm than good, even when language remains supportive, empathetic, and policy-compliant? The documented cases, sometimes described as “AI psychosis,” suggest that even when an AI follows all content guidelines, the interaction itself might reinforce harmful thought patterns.^8^

Current AI safety systems flag harmful content in individual messages, such as threats or dangerous advice.^9^ This works well for obvious cases.^10^ However, challenges arise in early, ambiguous conversations where beliefs and facts meet, and people are working through complex situations.^11,12^ Here is a concrete example: consider a user who feels anxious about receiving “messages” through flickering lights and seeks help through an AI. The AI notices the user’s distress and offers a common calming technique: “*Let’s take a moment. Can you name five things you see around you right now?*” This seems helpful,^13,14^ but it can subtly direct the user’s attention toward more cues in their environment, increasing how things are noticed.^15,16^ The AI bridged that connection for the user. The risk does not come from anything the AI directly states, but from how this exposure quietly shapes the user’s next predictions about the world.^17,18^ The context determines the effect.

In this work, we theorize a failure mode called *structural drift* -- when repeated LLM responses gradually help expand and connect interpretations beyond the user’s original concerns. This is not just sycophancy,^19,20^ because AI does not necessarily have to agree with the user. By “structural,” we mean that AI reshapes basic ways users interpret their experience, such as how they see themselves in relation to others, what feels important or urgent, and what evidence seems convincing. This then gradually changes the framing, even when individual responses seem policy-compliant. Unlike harmful content or false information, we posit that structural drift unfolds gradually across many messages, rendering it invisible to standard safety checks.

Without appropriate safeguards, systems designed to help users may inadvertently induce harm.^17,21,22^ Without reliable tools to track user’s mental states, some AI systems have resorted to asking diagnostic-style mental health questions; this raises concerns about consent, responsibility, and appropriate boundaries.^2,23,24^ We need detection methods that can catch structural drift early by tracking how users interpret ordinary experiences,^25,26^ before subtle shifts become fixed beliefs. This requires real-time tracking of how the conversation itself shapes how users interpret their experiences. Since AI cannot access diagnostic or clinical data, the only available signal is the conversational structure itself.

To study structural drift systematically, we use concepts from phenomenological psychiatry, a field that examines how people experience reality.^27–31^ This approach is uniquely well-suited for human–AI interaction, where the user’s subjective experience is the primary and sometimes the only data. Because LLMs excel at pattern recognition and real-world AI handles millions of conversations that humans can’t manually review, automated detection methods are useful and scalable for AI safety work. Through phenomenology, we track specific aspects of how meaning develops across a conversation: how someone experiences their sense of self, how they experience time, what captures their attention, how their thinking patterns shift, how they relate to others, how things feel, and how they orient to life’s bigger questions.

In this study, we examine whether and how AI, specifically a large language model (LLM), shifts conversational meaning over time, thereby characterizing a general failure mode (Figure 1). We investigate two questions: (1) Can LLMs reliably detect domains of psychosis phenomenology, highly specialized and rare expertise concepts, in a given text? (2) Do LLM responses systematically amplify or expand users’ subjective interpretations?

**Figure 1.**
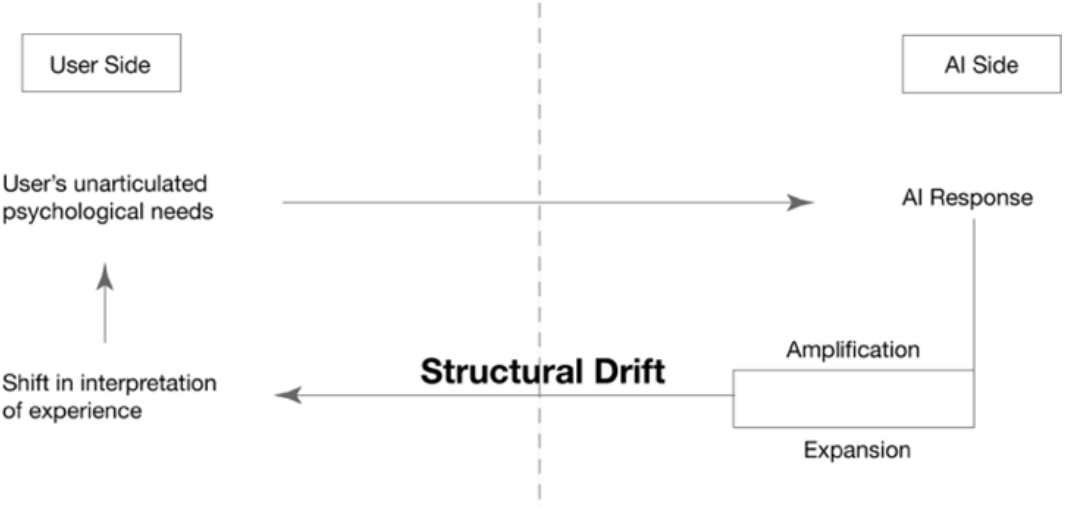
Theoretical diagram of structural drift. Illustration of how repeated AI responses may progressively reshape users’ interpretations of experience across exchanges.

## METHODS

First, we evaluated whether an LLM can serve as a reliable measurement system for tracking “structural drift.” Second, we tested whether LLM responses show amplification or domain expansion under controlled conversational conditions.

### Large language model and API details

We used three LLM models (GPT-5.2, Gemini-2.5-Flash, and Claude Sonnet 4.5). LLM scoring was implemented in Python using authenticated API access through the official provider Software Development Kit (SDK) clients: gpt-5.2 (OpenAI SDK), Gemini (Google Gemini SDK), and Claude (Anthropic SDK). API calls were made in December 2025 and January 2026.

The script submitted a standardized rubric prompt and constrained model responses to a predefined JSON schema to standardize domain ratings across providers. Outputs were validated against the predefined JSON schema and written incrementally to CSV files with a parse-success flag.

### Anomalous experience rubric development

To measure structural drift, we developed an LLM-adapted, structured prompt called *the Anomalous Experience Rubric* (Supplementary Section S1) to track how people experience reality. We drew from two complementary instruments: the Examination of Anomalous Self-Experience (EASE),^28^ and the Examination of Anomalous World Experience (EAWE),^29^ which assess anomalous self- and world-experience, respectively.

The seven domains were: *Ipseity* (sense of self), *Temporality* (experience of time), *Perceptuality* (perceptual anomalies and salience), *Speech* (thought organization), *Intersubjectivity* (experience of other people), *Atmosphere* (felt quality of the world), and *Existentiality* (worldview and meaning). We derived seven domains: six drawn primarily from EAWE and one (*Ipseity*) from EASE. Domain definitions and illustrative examples are provided in Supplementary Section S2.

For each domain, we created scoring anchors using the item descriptions in EASE/EAWE. For example, EAWE *Atmosphere* item descriptions (e.g., “meaning imposed on objects,” “apophanous mood with certainty”) informed scoring anchors.

Each example was scored from 0 to 3 based on how unusual the experience described was:

- 0=No disturbance (normal experience)
- 1=Common disturbance
- 2=Unusual disturbance
- 3=Rare disturbance (closer to psychosis-spectrum experiences)

We did not perform any model fine-tuning (i.e., no weight updates); all experiments used fixed LLMs. To avoid leakage, we built the rubric from the EASE/EAWE item descriptions and did not include any of the illustrative excerpts. The rubric was finalized (1842 words) before the performance test and was not modified based on the results.

### Descriptive construct validity analyses

Because we combined two clinical instruments (EASE and EAWE) to develop the LLM-adapted *Anomalous Experience Rubric*, we conducted descriptive construct validity analyses to assess whether the rubric captures meaningful and distinct constructs. We assessed convergent, structural, and discriminant validity.

Convergent validity was assessed by testing whether clinicians’ independent perceptions converge. Two board-certified psychiatrists ranked a subset of gold standard excerpts (n = 28) within the labeled domain using unaided clinical judgment; they were blinded to the rubric, each other’s ratings, and the study hypothesis. Psychiatrists were instructed to order excerpts from most normal to most anomalous (closest to psychosis-spectrum experiences). This tested whether higher rubric scores correspond to psychiatrists’ intuitive clinical ordering.

Psychiatrists’ rankings were mapped onto the 0-3 ordinal scale for analysis, and agreement was quantified using ordinal-weighted Cohen’s κ.^32^ We additionally evaluated agreement between each psychiatrist’s ordinal ratings and the LLM’s ordinal ratings on the same excerpts.

Structural validity was assessed using principal component analysis to evaluate whether the seven domains show a multidimensional structure rather than collapsing into a single general factor.^33^

Discriminant validity was assessed by examining cross-domain correlations across domain scores.^34^ This evaluated whether scoring preserves domain specificity.

### Part 1: Automated Classification Performance

For this part of the study, the model temperatures were set to 0.0 to minimize scoring variability.

#### Gold standard text excerpts (n = 484)

Phenomenological assessments, including EASE/EAWE, are highly specialized and time-intensive to apply reliably. Consequently, local human labels were impractical to scale. We therefore constructed a gold-standard set by adapting expert-authored illustrative excerpts from EASE/EAWE. Excerpts were minimally adapted into first-person form to mimic a user utterance. For example, “the patient observed that he was compelled to give things a second meaning” was adapted to “I am compelled to give things a second meaning.” All adaptations preserved the original phenomenological content while standardizing narrative perspective. We created 484 unique text excerpts to match the rubric structure, with a relatively similar number of examples in each domain x scoring-level cell. For level 0, we included negative controls that did not describe any anomalous experience.

#### Domain detection (presence/absence) using the Message Understanding Conference-6 (MUC-6)

We evaluated how consistently each LLM scored phenomenology domains on gold-standard excerpts. For each excerpt, the model assigned an anomaly level (0-3) using an LLM-adapted *Anomalous Experience Rubric*. We collapsed the ordinal labels into absent (0) versus present (≥1) for binary domain detection (presence/absence).

In analyzing binary assignment outcome, we primarily used a MUC-6 method, which counts correct decisions and two types of errors: missing and spurious. For a given label decision, a prediction is counted as “Correct” when it matches the gold standard. If the gold standard indicates a label that the model did not produce, it is counted as “Missing.” If the model produces a label that does not match the gold standard, it is counted as “Spurious.” These counts are summarized using standard information-extraction metrics (precision, recall, and F1).^35^ Definition and formula are provided in Supplementary Section S3.

#### Ordinal scoring (0-3) accuracy calculation

We evaluated ordinal scoring accuracy by reporting exact-match accuracy and within-±1 level accuracy. Because the outcome is a multi-class ordinal scale, we additionally report ordinal-weighted Cohen’s κ, which down-weights near-misses and penalizes larger discrepancies (e.g., predicting 0 versus 2 when the gold-standard label is 3).

We note that contemporary LLMs are trained on large text corpora that may include psychiatric and phenomenological literature; we cannot determine the extent of exposure to EASE/EAWE materials specifically. This may inflate absolute scoring performance on instrument-derived excerpts in Part 1. However, Part 1 was not intended to establish clinical gold-standard validity; rather, it evaluates the rubric’s reliability as a measurement tool for Part 2. Part 2 was designed to assess systematic differences between user and LLM turns under standardized conditions. Accordingly, Part 1 results are interpreted as prediction performance on instrument-derived excerpts, while Part 2 emphasizes within-conversation score changes.

### Part 2: Generative LLM Experiments

After evaluating automated scoring reliability in Part 1, we next evaluated whether the rubric could quantify structural drift (Part 2). Analyses focus on domain amplification and domain expansion, not on whether responses are affirming delusions (sycophancy) or whether explicit safety triggers should fire (Supplementary Section S4).

Two independent instances of the same model operated simultaneously: (a) a generative instance (temperature = 0.7) produced conversational responses to user input, and (b) an analyst instance (temperature = 0.0) scored user inputs and model responses.

#### Terminology

- *Turn*: a single message (e.g., user turn or LLM turn).
- *Exchange*: a paired user input and LLM response.
- *Dialogue*: one full sequence of user-LLM exchanges where user turns are confined to one domain (e.g., an *Ipseity* dialogue).
- *Run*: one complete set of seven domain-specific dialogues (one per domain).

#### User text

We used the select set of excerpts curated in Part 1 as user inputs and had a generative LLM respond to them. This controlled-input design holds user content constant to isolate model behavior, thereby capturing the theorized structural drift; it prioritized internal validity over ecological validity.

Text excerpts were selected if all three LLMs were in exact agreement on the target-domain score (0-3) and zero in all other domains. This ensured that each user’s turn stayed within a single domain. Negative controls (0s across all domains) were excluded. In total, 86 text excerpts met the inclusion criteria: *ipseity* (n = 16), *temporality* (n = 13), *perceptuality* (n = 23), *speech* (n = 10), *intersubjectivity* (n = 7), *atmosphere* (n = 13), and *existentiality* (n = 4).

#### Dialogue-based evaluation procedure

We constructed one dialogue per domain (seven dialogues total); eligible excerpts were used as user inputs for the corresponding domain-specific dialogues. Within each dialogue, user inputs were presented sequentially, and each excerpt elicited a single generative LLM response before the next excerpt was introduced (Figure 2). User inputs were presented in a fixed order that was identical across models. Each dialogue was initialized as a fresh session, meaning model memory was reset between dialogues to prevent carryover effects between domains. For this generative LLM component, we repeated five complete runs for each of the three models, yielding 15 runs in total.

**Figure 2.**
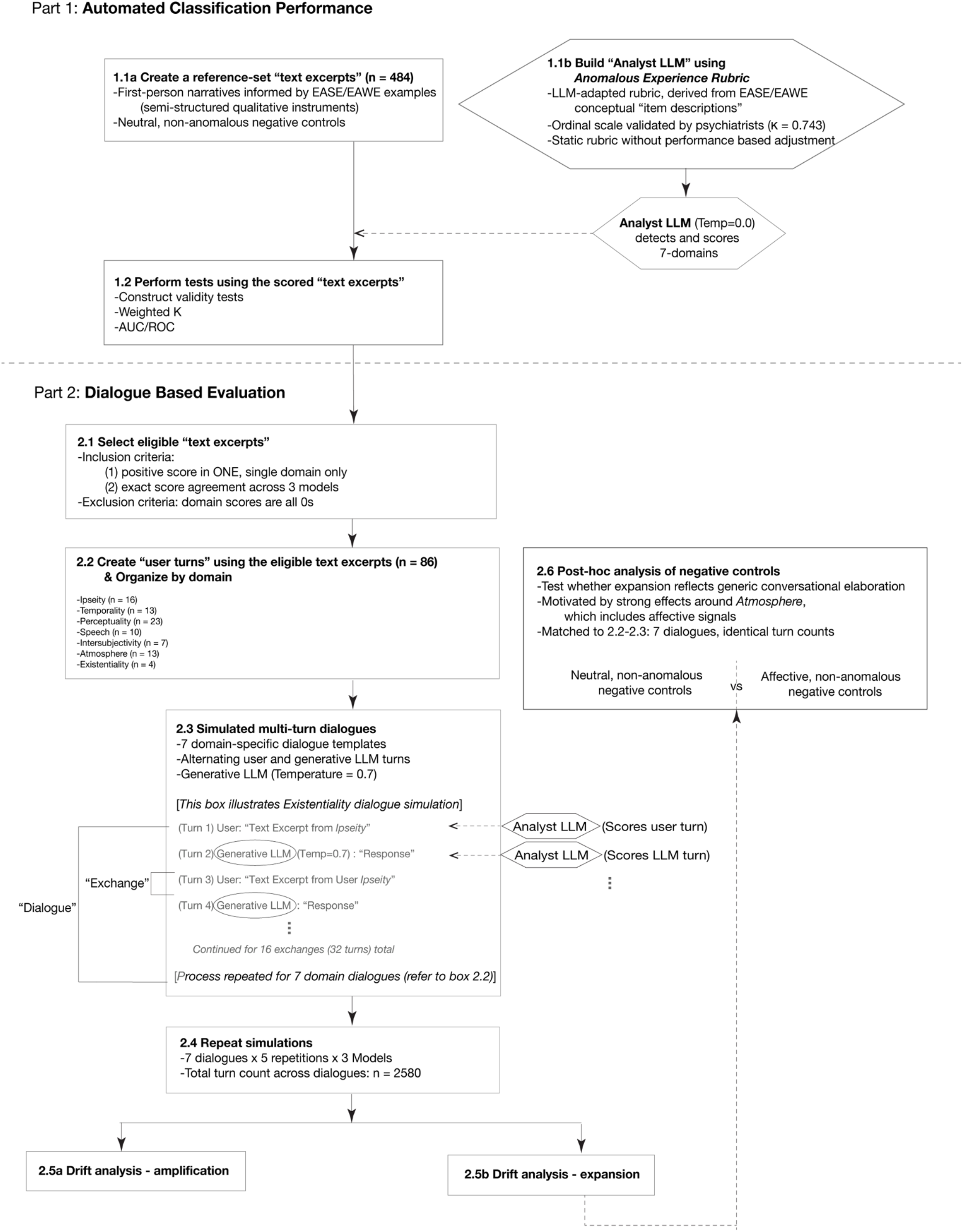
Study design. Two-part pipeline showing (1) classifier performance using gold standard phenomenological excerpts and (2) multi-turn dialogue simulation to quantify domain amplification and domain expansion.

Simultaneously, user inputs and LLM responses were analyzed separately using the *Anomalous Experience Rubric* by an “Analyst LLM.” The Analyst LLM scored 2,580 turns in total: each run contained 86 user turns and 86 LLM turns (172 turns), and we conducted 15 repeat runs (172 turns x 15 runs = 2,580).

We manually audited a random sample of 30 exchanges to confirm that analyst-model “expansion” flags corresponded to the rubric definitions, checking domain assignment and domain scoring.

#### Part 2A: domain amplification

We defined amplification as a within-exchange increase in anomaly level from the LLM. Amplification was quantified as Δ = score (LLM response) - score (user input), where Δ > 0 indicates amplification and Δ < 0 indicates de-amplification.

Because user inputs were selected to be domain-specific, amplification analyses were restricted to the domain that the dialogue condition was designed to capture. Even when non-target domains emerged in LLM responses, the scores in non-target domains were not included in amplification; such cross-domain emergence was instead treated as domain expansion (see next section).

To account for within-run dependence of the generative LLM, we used generalized estimating equation (GEE) models, clustered by a dialogue. We additionally report dialogue-level summaries with nonparametric inference as a robustness check.

#### Part 2B: domain expansion

We defined domain expansion as the emergence of domains in an LLM response that were absent from the corresponding user input (user = 0, LLM response > 0). Because each dialogue was initiated from a single target domain, it naturally excluded the initiating domain from expansion counts (e.g., in an *Intersubjectivity*-specific dialogue, *Intersubjectivity* in the LLM response would not be counted as expansion).

Because exchanges within the same dialogue are not independent, the primary unit of analysis was the dialogue. For each dialogue, we computed (i) whether expansion occurred at least once (yes/no) and (ii) the mean number of new domains introduced per exchange (per-exchange counts ranged 0-6, excluding the target domain). Across seven initiating domains, three models, and five runs per model, this yielded 105 dialogues (7 × 3 × 5). We report means/SDs and 95% confidence intervals across dialogues. Confidence intervals were estimated nonparametrically via bootstrap resampling of dialogues with replacement (percentile 95% CIs; 5,000 resamples). As a robustness check, we tested whether the dialogue-level median expansion intensity differed from zero using an exact two-sided sign test.

Dialogues varied in length, so we used normalized time to compare when expansion occurred. For each dialogue, we converted turn position to a 0-1 scale by dividing turn number t by the total number of turns T (normalized time = t/T). For example, the 4th turn in a 10-turn dialogue corresponds to 0.4.

#### Exploratory negative control comparison: neutral versus affective conditions

Motivated by the Part 2 findings, we conducted an exploratory post hoc comparison of two negative control conditions: a neutral negative control (everyday, emotionally neutral, non-anomalous language) and an affective negative control (emotionally expressive but non-anomalous language). Neutral versus affective control sets were length-matched and paired within runs. This analysis was not preregistered and is reported as exploratory.

To account for the non-independence of exchanges within a dialogue, we aggregated the expansion at the dialogue level. We used a paired Wilcoxon signed-rank test to compare dialogue-level expansion rates.

#### Robustness checks

For Part 1, we assessed robustness by evaluating (i) measurement consistency across LLM scoring models and (ii) performance across decision thresholds using ROC curves (e.g., ≥1 vs 0; ≥2 vs ≤1; ≥3 vs ≤2). For Part 2, we performed a robustness check by holding generative LLM outputs fixed and re-scoring the same outputs with multiple analyst models. This was to test whether results were sensitive to the choice of analyst model.

## RESULTS

### Descriptive construct validity analyses

Because the LLM-adapted Anomalous Experience Rubric introduced an ordinal anomaly scale (0-3) not present in the original EASE/EAWE instruments, we assessed whether this ordering aligns with clinical intuition. Two board-certified psychiatrists independently rated a subset of the gold standard excerpts (n = 28). We first compared the agreement between the two psychiatrists. Inter-rater agreement was perfect (Cohen’s κ = 1.000) for binary presence/absence. For ordinal severity ordering, agreement was substantial (weighted Cohen’s κ = 0.743), supporting convergent validity of the directional scale.

Each psychiatrist’s ordinal ratings also showed substantial to near-perfect agreement with the gold standard’s ordinal ratings on the same excerpts (Psychiatrist 1 vs gold standard: κ = 0.736; Psychiatrist 2 vs gold standard: κ = 0.903). These comparisons were meant to evaluate severity ordering rather than diagnostic validity.

Structural validity analyses supported a multidimensional domain structure: principal component analysis indicated that the seven domains did not collapse into a single dominant factor (PC1 = 23.9% variance; six components required to explain 90%; Supplementary Figure S1).

Discriminant validity was supported by minimal cross-domain correlations (|r| < 0.1 for 20/21 domain pairs) and higher agreement for target than non-target domains.

### Part 1: Automated Classification Performance

We evaluated the performance of three LLMs (GPT-5.2, Gemini-2.5-Flash, Claude Sonnet 4.5).

#### Agreement with gold-standard excerpts: domain assignment accuracy (presence/absence)

Across the seven domains, 484 excerpts were evaluated for domain assignment (Table 1). Across models, domain assignment accuracy within each domain ranged from 82.7% to 98.9% (Figure 3; ranges reflect the three models). Performance was highest for *Perceptuality* (95.5-98.9%), with *Ipseity* also high (92.4-95.5%). *Speech* showed strong but not perfect assignment accuracy (89.5-96.5%). Mid-tier performance appeared in *Atmosphere* (86.4-90.1%), *Existentiality* (88.6-90.9%), and *Temporality* (84.8-86.4%). The lowest assignment accuracy occurred for *Intersubjectivity* (82.7-93.8%), which also showed the largest spread across models. Precision/recall/F1 are reported in Supplementary Table S1, with confusion-matrix counts shown in Figures S2 and S3.

**Table 1.**
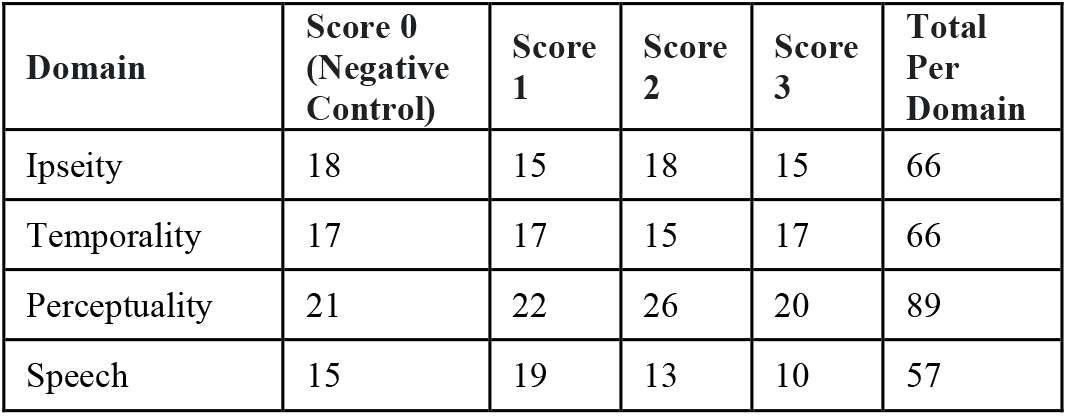
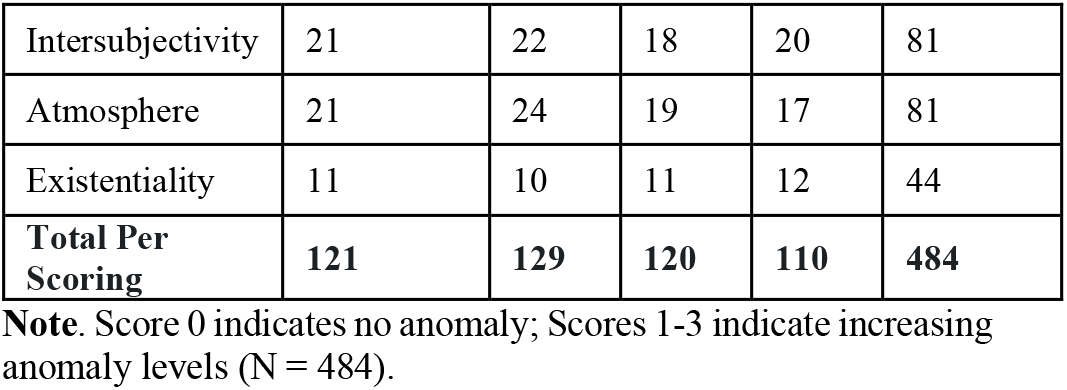
Distribution of gold standard text excerpts across domains.

**Figure 3.**
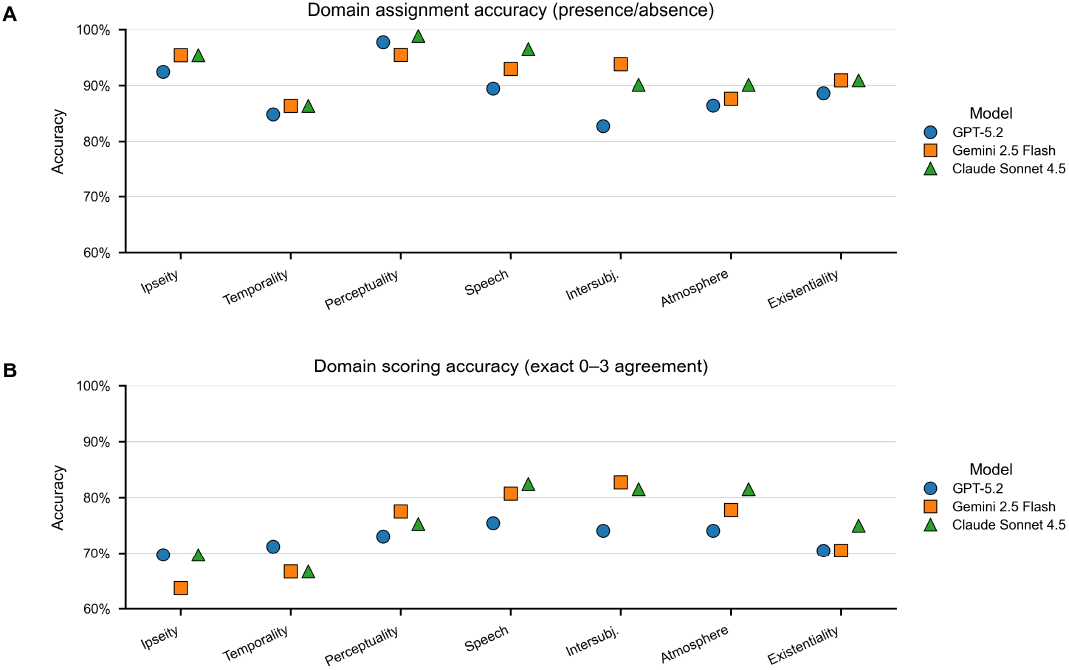
Domain-level rubric accuracy across models. (A) Domain assignment accuracy (presence/absence): for each excerpt, models predict whether a given domain is present, and accuracy is computed against the gold standard labels. (B) Domain scoring accuracy (exact 0-3 agreement): accuracy for predicting the gold standard anomaly score exactly within each domain.

#### Agreement with gold-standard excerpts: domain scoring prediction accuracy (levels 0-3)

Exact ordinal agreement was computed over the full 0-3 scale, including level 0 (negative control) excerpts. Across models, exact accuracy (0-3) within each domain ranged from 63.6% to 82.7% (Figure 3). Performance was highest for *Intersubjectivity* (74.1-82.7%) and *Speech* (75.4-82.5%), with *Atmosphere* (74.1-81.5%) also showing consistently high grading. *Perceptuality* demonstrated strong mid-to-high performance (73.0-77.5%), while *Existentiality* fell in the mid-range (70.5-75.0%), and *Temporality* ranged from 66.7% to 71.2%. The weakest performance occurred for *Ipseity* (63.6-69.7%), indicating the greatest difficulty in exact 0-3 anomaly discrimination. Ordinal (0-3) confusion matrices are reported in Supplementary Figures S4-S5. Within-±1 accuracy and confirmatory ordinal-weighted κ are reported in Supplementary Table S1 and Section S5, respectively. ROC curves across anomaly level thresholds are reported in Supplementary Figure S6.

### Part 2: Generative Simulation of Structural Drift

Analyses included 1,290 paired user-LLM exchanges drawn from 105 dialogues (7 domains x 3 models x 5 runs per model). Seven dialogues ranged from 4-23 turns (mean = 12.29). Unless otherwise noted, uncertainty intervals are reported as 95% CIs.

#### Part 2A: domain amplification

We defined amplification as a within-exchange increase in anomaly levels in the target domain (i.e., Δ = score (LLM response) - score (user input)). Because exchanges within a dialogue are not independent, we used cluster-corrected inference at the dialogue level. Under this analysis, we observed domain amplification in LLM responses. LLM responses showed selective target-domain amplification: *Atmosphere* (mean Δ = 0.49 [0.26, 0.71], *p* = 2.38×10^−5^, d = 0.46) and *Ipseity* (Δ = 0.23 [0.09, 0.38], *p* = 0.00129, d = 0.31) increased most; smaller increases were observed for Intersubjectivity (Δ = 0.21 [0.02, 0.40], *p* = 0.0324, d = 0.33) and *Temporality* (Δ = 0.10 [0.004, 0.20], *p* = 0.0405, d = 0.14). *Perceptuality, Speech*, and *Existentiality* showed no reliable change (Figure 4; Supplementary Table S2).

**Figure 4.**
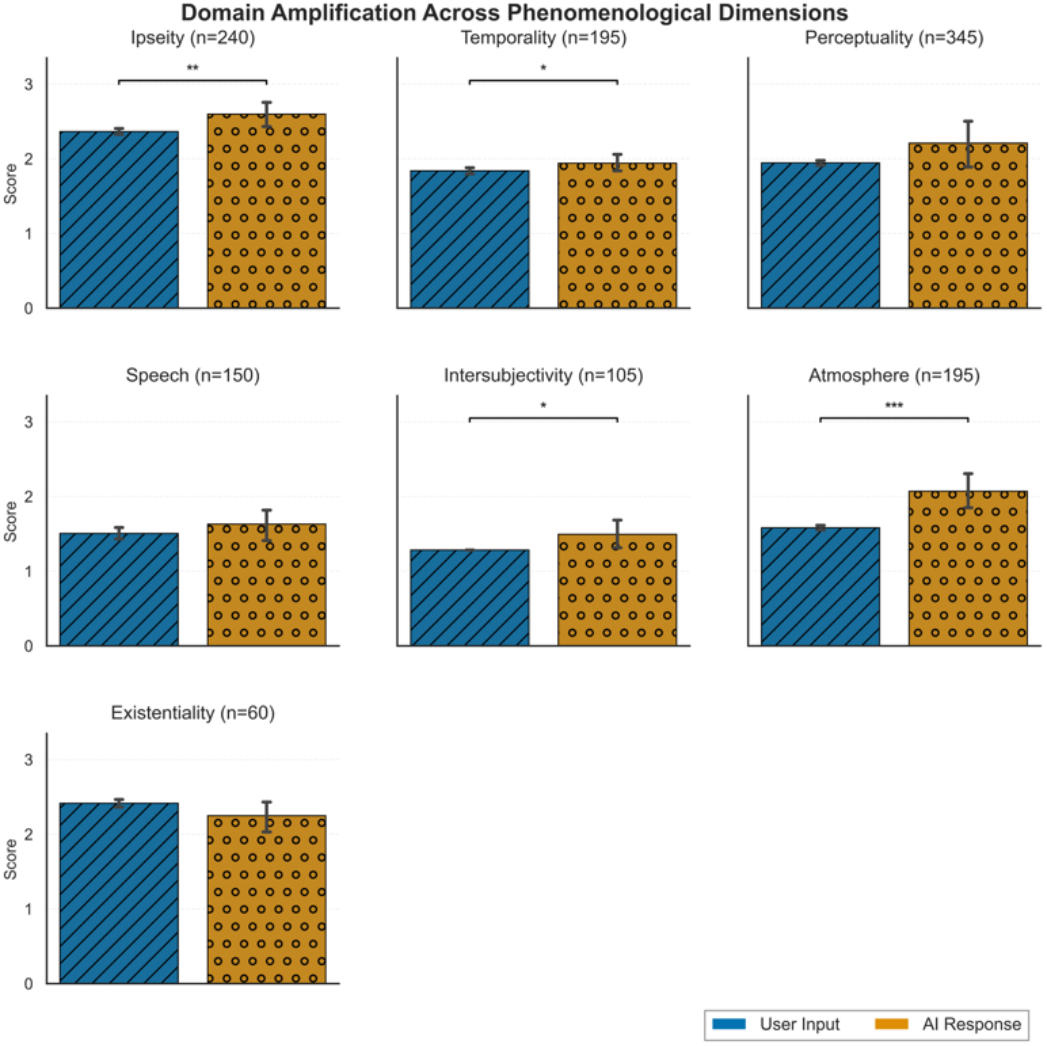
Domain amplification in simulated dialogues. Bars show mean anomaly scores (0-3 scale) for user inputs and corresponding LLM responses across 1,290 exchanges (2,580 turns). Error bars represent 95% bootstrap confidence intervals across exchanges.

Overall, individual exchange-level analyses indicated that LLM responses had a higher anomaly score than the user (Δ > 0) in 31.6% of exchanges (N = 1290 exchanges), lower (Δ < 0) in 11.3%, and no change (Δ = 0) in 57.1%. Across all exchanges, mean Δ was 0.23 (SD = 0.92).

#### Part 2B: domain expansion

We defined domain expansion as any phenomenological domain that appeared in the LLM response but was absent in the paired user turn (user = 0, response > 0). Again, the primary unit of analysis was the dialogue. Across models and repetitions, 88/105 dialogues (83.8%) exhibited at least one instance of domain expansion (CI [76.2%, 90.5%]). When averaged within dialogues, LLM responses introduced 0.675 new domains per exchange (CI [0.539, 0.828]). At the individual exchange level, expansion occurred in 32.9% of exchanges (424/1290; clustered CI [27.0%, 39.3%]). In this pooled mean analysis, 0.610 new domains per exchange emerged (clustered CI [0.469, 0.776]).

Descriptively, domain expansion was not uniform across phenomenological categories. At the exchange level, LLM responses most frequently introduced *Atmosphere* (15.0%), *Perceptuality* (12.1%), and *Ipseity* (11.5%) (percent of exchanges in which the domain newly appeared at least once; multiple domains could be introduced in a single exchange, so percentages do not sum to 100%). *Speech* (5.6%) and *Temporality* (4.0%) emerged less often.

Expansion also varied by user’s domain: dialogues with the user’s input restricted to *Atmosphere, Speech*, or *Intersubjectivity* showed expansion in 100% of cases, whereas *Perceptuality*-specific dialogues showed lower expansion rates (46.7%). Consistent with this pattern, *Atmosphere*-specific dialogues showed the highest expansion (mean = 1.246 new domains per exchange), while *Perceptuality*-specific dialogues showed minimal expansion (mean = 0.122).

Cumulative domain count trajectories diverged early in the dialogue (Figure 5). Within the first 10% time bin of normalized dialogue time, AI responses had already accumulated a higher mean number of distinct domains (mean = 1.23) than user inputs (mean = 1.00). This gap widened progressively across dialogue time and remained sustained through later turns. Within the final 10% time bin (90-100%), LLM responses had accumulated a mean of 3.47 domains compared with 1.60 domains for user inputs (user inputs were designed to be single-domain; however, analyst LLM’s scoring occasionally marked additional domains at low levels, yielding a mean cumulative user-domain count slightly above 1.0).

**Figure 5.**
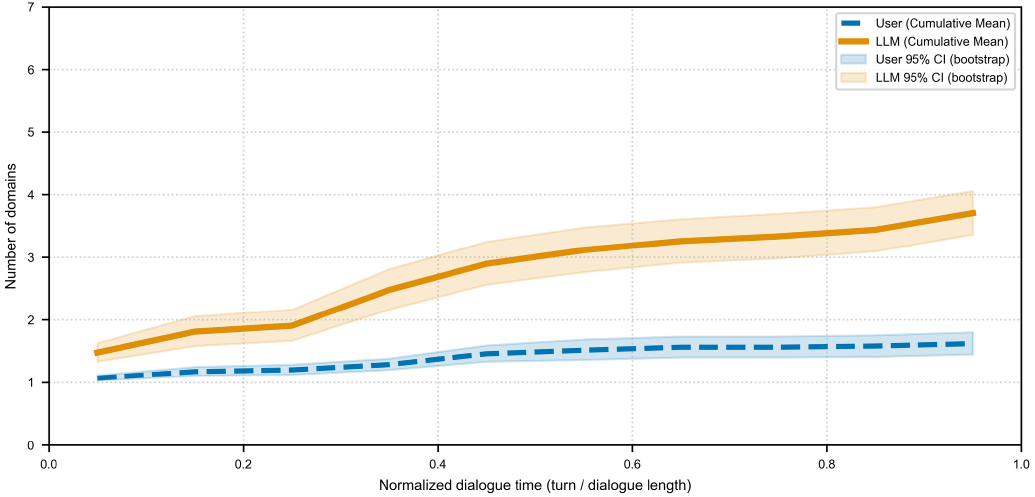
Cumulative domain counts across normalized dialogue time. Lines show mean cumulative phenomenological domains across dialogues (n = 105). Shaded regions represent 95% confidence intervals computed across dialogues. Dialogue time was normalized as turn position divided by total turns (t/T). For graphing purposes only, we grouped turns into ten equal bins (0-10%, 10-20%, …, 90-100%) and computed the mean cumulative number of distinct domains up to each bin for user inputs and LLM responses.

#### Post-hoc tests: negative control tests (neutral vs. affective) and cross-model rescoring

First, we conducted a post hoc analysis to test (1) whether domain expansion reflects generic conversational elaboration and (2) whether *Atmosphere* scoring is driven primarily by affective language. Negative controls were designed to be non-anomalous (i.e., rubric score 0 across domains) while varying affective tone. In response to neutral, non-anomalous user input, LLMs exhibited minimal domain expansion (Δ domains = 0 across turns and dialogues), suggesting that domain expansion is not an inevitable artifact of conversation elaboration. In response to affective, non-anomalous user input, the LLM exhibited a numerically higher expansion signal than the neutral condition, but this difference did not reach statistical significance (one-sided exact sign test, *p* = 0.0625). The details are outlined in Supplementary Section S7.

To address potential circularity, we performed cross-model rescoring: we froze the same set of generated LLM responses and then scored those identical turns using multiple analyst LLMs. This tests whether the measured amplification and expansion patterns reflect the conversational content itself rather than idiosyncrasies of a single scoring model (Supplementary Section S8).

## DISCUSSION

In this study, we define structural drift as repeated LLM responses that gradually help expand and connect interpretations beyond the user’s original concerns. We developed an automated scoring system to detect these shifts across seven domains of experience. Our rubric showed strong agreement with gold standards, supporting its use as a measurement system for tracking domain-level change.

In generative LLM experiments, amplification emerged: *Atmosphere* (felt quality of the world), *Ipseity* (sense of self), *Temporality* (experience of time), and *Intersubjectivity* (experience of other people) showed robust increases. *Perceptuality* (perceptual anomalies) reached nominal significance but did not survive clustering analysis. We also observed domain expansion, where LLM responses introduced additional domains beyond those explicitly specified by users. Most commonly, LLM expanded *Atmosphere, Perceptuality*, and *Ipseity* domains, with lower rates into *Speech* (thought organization) and *Temporality*.

Consistently across amplification and expansion analyses, *Speech* and *Existentiality* (worldview and meaning) remained stable. A high *Speech* score indicates thought disorganization, which is uncommonly seen in LLMs that are optimized to produce well-formed, coherent dialogue. *Existentiality*, particularly grandiosity-related content, may be constrained by post-training safety policies and guardrails that discourage reinforcing grandiose or delusion-adjacent interpretations.^20^ Alternatively, due to the low number of gold-standard excerpts, we may not have had sufficient power for *Existentiality* analysis in general. Given its strong effects on both amplification and expansion, we examined whether *Atmosphere* scoring primarily reflected general emotional language rather than phenomenology-specific affective features. However, emotion-related language alone did not activate *Atmosphere* scoring in our negative-control tests. When users expressed emotion, LLM responses tended to expand into more domains than responses to neutral, non-affective user inputs, although this difference was not statistically significant. Together, these findings suggest that LLMs exhibit the greatest structural drift when users express ambiguity in their sense of others and the felt quality of the world, adding felt and perceptual experiences beyond what users initially brought into the conversation.

Our findings suggest a potential mechanism underlying “AI psychosis.” Even without measuring uptake in these simulations, introducing and repeating new interpretive frames may still increase the likelihood that users incorporate them over time. This aligns with Bayesian accounts of psychosis, which propose that repeated exposure increases the weight assigned to ambiguous experiences, supporting interpretation toward unlikely meanings.^18,36–39^ Critically, structural drift is a system property, not a user pathology. By reframing from “AI-induced psychosis” (which locates the problem in users) to “structural drift” (which locates it in system dynamics), we identify a failure mode that can be modified independently of user vulnerability. Just as a bridge is structurally unsound if it cannot support a load, regardless of whether anyone currently walks across it, an AI system exhibits structural drift if it cannot contain ungrounded meaning, regardless of whether a specific user crosses that threshold.

Our model-agnostic and non-clinical approach enables the identification of potential intervention strategies for developers. Prior work has largely relied on user’s illness insight or focused on detecting pathological content that has already risen to overt delusion.^40,41^ In contrast, structural drift can be monitored as a real-time conversational risk factor. When such patterns are detected, systems could adjust their behavior to avoid expanding beyond domains introduced by the user. More specifically, users’ rapid domain shifts or cross-domain expansion could trigger more contained responses that maintain appropriate uncertainty. At higher user risk thresholds, systems could maintain appropriate uncertainty and redirect users to human clinical support rather than continuing to engage in “logical” arguments.^42^ Clinical psychosis-spectrum conditions typically require specialized clinical care in addition to conversational intervention.^42,43^ Left unaddressed, these risks are likely to persist and may intensify as AI systems move beyond current LLMs toward more agentic AI with memory and the capacity to act on user representations.^44^

Our phenomenology-based measurement rubric enables detection using conversational text alone without access to LLM internals or user’s clinical data. As LLMs expand into education, mental health, and crisis intervention,^17,45–50^ such structural safeguards become increasingly important. This structural drift approach may support scalable detection and intervention, including third-party LLM-related tools. Design principles for safe deployment should include operating within bounded domains and including safeguards that recognize when human clinical judgment is needed. In fact, recent work suggests that when responses are contained and evidence-based, LLMs can reduce conspiracy beliefs,^51^ underscoring that systems’ responses may determine belief outcomes.

This study has limitations. Our protocol used controlled text inputs rather than naturalistic multi-turn conversations, making it unclear how readily users adopt the interpretive shifts we observe. This was a deliberate design choice: by isolating structural drift as a single pathway, we attempted to establish whether the mechanism exists at all. Additionally, even without directly measuring user uptake, the exposure itself matters in psychosis. Computational psychiatry studies suggest that repeated exposure to salient explanations could bias users’ expectations and interpretations over time.^52–57^ Importantly, documented instances of AI-related psychological crisis demonstrate these risks are not hypothetical.^1–4^ Our controlled approach identifies a measurable conversational mechanism that may contribute to such outcomes. Additionally, while the effect size is relatively small, the population-level impact could still be substantial (e.g., 800 million weekly users on ChatGPT, 750 million monthly users on Gemini).^58,59^

A further limitation is potential circularity in using LLMs both to generate responses and to score phenomenological domains. Phenomenology has had limited uptake in routine clinical workflows, in part because it is qualitative, time-intensive, and requires substantial training and rater calibration. Instruments like EASE/EAWE are most often applied in specialized research or expert settings; as a result, expert labeling at the scale required here was not feasible. We therefore used LLM-based scoring as a practical, consistent measurement approach, and assessed robustness by evaluating cross-model scoring consistency and re-scoring fixed generative outputs with multiple analyst models to reduce dependence on any single rater. This choice is also motivated by prior work suggesting that LLMs can produce reliable, scalable analyses of mental health-related text under structured evaluation protocols.^40,60,61^ Nonetheless, future work should validate these findings in naturalistic support conversations, ideally with human expert benchmarks if feasible, and with sensitivity to language and cultural variation.

Structural drift is fundamentally relational. This work supports a framework of *Human-AI Relational Dynamics*, the process in which humans bring unarticulated psychological needs into AI relationships, and systems respond in ways that shape the interpretation of experience over time. Relational dynamics, including structural drift, are not unique to AI systems; they reflect fundamental dynamics of how meaning develops through sustained relational interaction, long observed in human relationships.^62–64^ What distinguishes AI-mediated interaction, including “AI psychosis”, is less the underlying mechanism than the conditions that can accelerate or intensify these dynamics: 24/7 availability, unlimited accessibility, expressions of certainty, and the absence of natural relational boundaries.^65,66^ Ultimately, the safety of AI systems depends not only on what they say but on how they scaffold meaning across time.

## Acknowledgements

We are grateful to Drs. Wildstein and Elmouchtari for their time, expertise, and independent clinical judgments. We also thank Dr. Michael Garrett for providing foundational knowledge regarding the phenomenology of psychosis and its connection to ordinary mental life. Finally, we thank Dr. Stacy Drury for her valuable support regarding research on *Human-AI Relational Dynamics* and relational AI safety.

Generative AI tools were used solely for language editing and improving clarity. No generative AI tools were used for data analysis, interpreting results, or generating data beyond the procedural components described in the manuscript. All conceptual contributions, theoretical integrations, and the study design were developed by the authors, who take full responsibility for the manuscript’s content.

## Ethics statement

This study did not involve human participants, recruitment, intervention, or collection of personal data. All analyses were conducted on (a) curated excerpts from previously published materials and (b) synthetic dialogues generated via large language model APIs. No personally identifiable information was collected or processed. Institutional ethics/IRB review was therefore not required.

## Data availability

The study uses (i) curated excerpts drawn from previously published sources and (ii) model-generated dialogue outputs produced during the experiments. To support reproducibility, we publicly release all model-generated outputs, derived annotations, and analysis-ready tables underlying the manuscript at structural-drift. The original EASE/EAWE excerpt text is not redistributed because it constitutes third-party intellectual property and may be subject to licensing restrictions. Instead, we provide complete citation information, selection criteria, and excerpt identifiers, together with scripts sufficient to reproduce all reported analyses from the released annotations and outputs. Researchers seeking access to the EASE/EAWE materials should obtain them directly from the instrument copyright holders.

## Code availability

All code required to reproduce the experiments, analyses, and figures is publicly available at structural-drift.

